# Evaluating algorithm Fairness in Predicting Health Service Use and Unmet Need Across Socioeconomic and Caste Subgroups: Evidence from Longitudinal Ageing Study in India

**DOI:** 10.1101/2025.09.09.25335385

**Authors:** John Tayu Lee, Vincent Cheng-Sheng Li, Toby Kai-Bo Shen, Valerie Tzu Ning Liu, Sheng Hui Hsu, Tzu-Pin Lu, Arokiasamy Perianayagam, Rifat Atun

## Abstract

**Introduction:** Persistent socioeconomic and caste inequalities in India drive disparities in healthcare access. Machine learning (ML) models offer promise for forecasting service use and unmet needs, but may perpetuate algorithmic bias against disadvantaged groups. We evaluated both performance and fairness of several ML algorithms across diverse caste and socioeconomic subgroups.

**Methods:** We used nationally representative data from India to develop machine learning models predicting outpatient care, hospitalization, and unmet healthcare need among older adults. We trained logistic regression, random forest, XGBoost, and LightGBM models using demographic, social, and health-related predictors. Synthetic Minority Oversampling Technique (SMOTE) was applied to address class imbalance. We assessed model performance using AUROC and evaluated fairness across caste and income subgroups. Fairness strategies included removing sensitive features (neutral models) and training stratified models within subgroups. We used SHapley Additive exPlanations (SHAP) to identify the most influential predictors across outcomes.

**Results:** Among 55,962 older adults in India, 53.4% had at least one outpatient visit, 6.5% were hospitalized, and 7.9% reported unmet healthcare needs. Model performance varied across outcomes and groups. The best-performing model (LightGBM) achieved AUROCs of 0.78 for unmet need, 0.76 for outpatient care, and 0.70 for hospitalization. Predictive accuracy was higher in the lowest socioeconomic group (MPCE 1, AUROC = 0.79) compared to the highest (MPCE 5, AUROC = 0.75). Removing sensitive predictors such as caste or income had minimal impact (change in AUROC <0.02), and subgroup-specific models led to mixed results, with only marginal improvement for Scheduled Castes (AUROC from 0.78 to 0.80). Including social and health determinants substantially improved model performance (e.g., hospitalization AUROC increased from 0.57 to 0.70). Top predictors included self-rated health, region, grip strength, and socioeconomic status. Balancing techniques like SMOTE did not meaningfully enhance performance.

**Conclusions:** Machine learning models can effectively predict healthcare use and unmet needs among older adults in India. Incorporating social and health determinants improves model accuracy, but eliminating bias requires structural changes beyond technical adjustments.

Fairness-aware model development and deployment are essential to ensure predictive tools contribute to more equitable healthcare systems.

**What is already known on this topic:**

- Machine learning (ML) has shown promise in predicting healthcare use and unmet need, particularly in high-income settings.
- Structural inequalities such as caste and income may influence healthcare access, but few ML studies have evaluated how these social factors affect model performance.
- Fairness concerns in ML are increasingly recognized, yet methods to assess or address them in low- and middle-income country (LMIC) settings remain limited.

**What this study adds:**

- This study evaluates ML model performance across caste and income subgroups using nationally representative data from older adults in India.
- It shows that model accuracy varies by subgroup, with better performance among Scheduled Castes and lower-income groups for predicting unmet need.
- Fairness interventions such as removing sensitive features or training stratified models offer limited benefit and do not fully resolve performance disparities.
- SHAP analysis identifies social and health determinants—especially self-rated health, caste, region, and income—as key drivers of predictions.

**How this study might affect research, practice or policy:**

- Encourages routine subgroup evaluation to ensure ML models do not exacerbate existing health inequities.
- Challenges the assumption that removing sensitive variables like caste or income improves fairness, emphasizing the need to address structural drivers directly.
- Supports the integration of social determinants into model development to enhance equity, transparency, and relevance in public health applications.

## INTRODUCTION

India, the most populous country in the world, has profound socioeconomic and caste disparities in healthcare access and outcomes ^1,2^. Caste groups include Scheduled Tribe (ST), Scheduled Caste (SC), Other Backward Class (OBC), and Others, with the first three being the most disadvantaged ones. These caste groups and the ones with lower socioeconomic backgrounds often experience significant barriers, including higher out-of-pocket expenditures^1,3^ and lower utilization rates of essential services ^4,5^. These inequities are compounded by systemic factors, including caste-based discrimination, economic inequities in income, and provision of inadequate healthcare infrastructure for lower caste groups, particularly in rural areas ^6,7^. Such challenges present critical obstacles to achieving Sustainable Development Goals (SDG) 3 (Good Health and Well-Being) and SDG 10 (Reduced Inequalities)^8^.

Machine learning approaches are increasingly employed in predictive health policy models to identify patterns in health-related data, allocate resources, and inform interventions ^9,10^. However, evidence highlights that these models are often prone to algorithmic biases, defined in relation to AI and health systems as “the instances when the application of an algorithm compounds existing inequities in socioeconomic status, race, ethnic background, religion, gender, disability or sexual orientation to amplify them and adversely impact inequities in health systems”^11^, particularly when applied to diverse and heterogeneous subgroups^12,13^. Such biases can result in unequal performance across populations, potentially perpetuating or exacerbating existing disparities ^14^. For instance, models may underdiagnose or misallocate resources to underserved populations, such as lower socioeconomic and caste groups, due to biased training data or flawed model design ^13,15^. Addressing these biases requires integrating equity-focused strategies throughout the model lifecycle, including data collection, model training, validation, and deployment, to ensure fair and effective healthcare interventions ^13,16,17^. In this study, we aim to predict three important key health service indicators as our study outcomes of interest, namely (i) outpatient care utilization, (ii) hospitalization, and (iii) unmet healthcare needs, across socioeconomic and caste groups in India and assess the fairness of model performance in terms of AUROCs across these subgroups.

Panel 1: Algorithmic Fairness in Machine Learning

Algorithmic fairness refers to the principle that machine learning models should perform equitably across different population groups—especially those historically disadvantaged. In health prediction, this means that model accuracy, sensitivity, and risk scores should be consistent across social subgroups such as caste, income, gender, or region.

There are several dimensions to model fairness:

1. Performance parity – The model should perform equally well across groups (e.g., similar accuracy, AUROC, sensitivity).
2. Equal opportunity – The model should provide similar true positive rates (e.g., detect unmet healthcare needs fairly in all groups).
3. Calibration – The predicted risk scores should mean the same thing across groups (e.g., a 70% predicted risk should have the same implication for a Scheduled Tribe woman as for a higher-income).

## METHODS

### Dataset

This study utilized cross-sectional data from the Longitudinal Ageing Study in India (LASI), a nationally representative survey collecting high-quality data from individuals aged 45 years and older. LASI is the most comprehensive ageing study in India, designed to provide extensive information on chronic health conditions and healthcare utilization. Data from the first wave of LASI, conducted in 2017/2018, were used for this analysis. The survey used a multistage stratified sampling design in both rural and urban areas to ensure national representativeness. Detailed caste data were incorporated into the weighting process to reflect India’s diverse population.

Initially, the survey included 72,250 older adults aged 45 years or older and their spouses, enrolled through multistage probability sampling. Due to incomplete biomarker module data from 6,350 individuals, the dataset was reduced to 64,867 participants. Further data cleaning and processing to address incomplete records resulted in a final analytical sample of 55,962 participants, representing 35 states and union territories.

LASI categorized caste groups into Scheduled Tribe (ST), Scheduled Caste (SC), Other Backward Class (OBC), and Others. Socio-economic groups were primarily defined using household consumption expenditure (MPCE) quintiles, with additional consideration given to educational attainment.

### Study Outcome Measures

We examined three primary outcomes within the past year of the survey: outpatient care utilization, hospitalization, and unmet healthcare needs. Outpatient care utilization was determined by whether participants reported receiving any healthcare services or consultations from a provider in the previous 12 months. Hospitalization was identified based on self-reported hospital admissions or overnight stays in a long-term care facility during the preceding 12 months; this was coded as a binary variable, with one or more hospital admissions treated as 1, and none treated as 0.

Unmet healthcare needs were assessed by determining whether participants had visited any medical facilities or providers in the past 12 months. Responses were coded in a binary format: individuals who reported zero consultations for reasons other than “did not get sick” were assigned a code of 1 (indicating unmet healthcare needs), while those who reported any consultations were assigned a code of 0 (indicating healthcare needs were met).

### Model Predictors

A total of 130 predictors, including comprehensive range of demographic, socioeconomic, health, lifestyle, and social factors to provide a holistic profile of participants: Demographic characteristics: Age, gender, marital status, caste, place of residence (rural or urban), religion, migration status, household size, and whether the household head was female.

Socioeconomic factors: Education level, working status, occupation, pension status (receipt of pension and pension amount), monthly per capita expenditure (MPCE), and home ownership.

Health and medical indicators: Chronic conditions (diabetes, cancer, chronic lung and heart diseases, stroke, bone or joint disorders, psychiatric conditions, high cholesterol, chronic renal disease, incontinence, and Benign Prostatic Hyperplasia [BPH]); vaccination history (influenza, pneumococcal, hepatitis B, typhoid, and diphtheria-tetanus); and infectious diseases (malaria, dengue, chikungunya, tuberculosis, and urinary tract infections). Functional and physical health: Activities of Daily Living (ADL), Instrumental Activities of Daily Living (IADL), cognitive score, depression, self-rated health, self-rated vision, pain, sleep quality, grip strength, use of aids (e.g., hearing aids, mobility aids), and denture usage. Anthropometric measures: Body Mass Index (BMI) categories (underweight, healthy weight, overweight, and obese) and waist circumference.

Lifestyle and behavioral factors: Tobacco use (smoking and smokeless), alcohol consumption, physical activity levels (types and intensity), food availability, and yoga practice. Social and family dynamics: Interaction with grandchildren, financial roles (providing or receiving financial support), family’s ability to perform daily activities, membership in organizations, experiences of discrimination, and role in household decision-making. Health insurance coverage: public and private.

### Data Preprocessing

We applied 1-hot encoding to preprocess categorical variable with more than 2 categories. All preprocessing steps, including handling missing data, variable encoding, and feature engineering, were documented and are available on GitHub. After preprocessing, the dataset was split into training (80%) and testing (20%) subsets. The training set was used to develop predictive models, while the testing set was reserved for evaluating model performance.

### Statistical Analysis and Modeling

We developed predictive models using multiple commonly used ML algorithms for structure health data: Random Forest, XGBoost, Gradient Boosting, LightGBM, Deep Neural Networks (DNN), and Fully Convolutional Networks (FCN).

#### Accuracy

assessed the algorithm’s performance by examining the area under the Receiver Operating Characteristic Curve (AUROC) and its 95% CI^18,19^, sensitivity (recall)^20^, specificity^20^, precision (positive predictive value)^20^, and F1 score (harmonic mean between sensitivity and precision)^20^. Receiver Operating Characteristic (ROC) curves and Area Under the Receiver Operating Characteristic Curve (AUROC) metrics were used to visualize and summarize model discrimination. Additional AUC comparison plots were generated to provide detailed insights into subgroup-specific model performance^21^.

#### Assessing Fairness

To examine the algorithms fairness. We conducted analysis based on caste and socioeconomic subgroups. We tested the following 3 approaches: first, we examined the performance of the full sample model when applied to each socioeconomic and caste subgroup to examine potential biases, recognized by the degree of differences in AUROC. Second, we generated neutral model, by removing the subgroup-sensitive predictive. Third, we generate stratified model that were specific to each subgroup. All analyses were done using Python programming language, version 3.11.11.

### Feature Importance and Subgroup Analysis

Feature importance, which quantifies the contribution of each predictor variable to the overall performance of a model, was analyzed using traditional statistical methods and SHAP (SHapley Additive exPlanations) values, derived from subgroup-specific models. Traditional methods quantified the overall contribution of predictors to model performance^22^, while SHAP values, grounded in cooperative game theory, attributed contributions to individual predictions. These methods enhanced model interpretability by identifying key factors influencing predictions^23^.

To assess the consistency of key features across subgroups, feature importance was examined within each subgroup, highlighting potential overlaps or differences in predictors among varying patient profiles. Statistical tests were applied to compare feature importance and model performance across subgroups, providing a deeper understanding of subgroup-specific dynamics^24^.

### Patient and Public Involvement

Patients and/or the public were not involved in the design, conduct, reporting, or dissemination plans of this research.

### Ethics

We used publicly available, deidentified data from the Longitudinal Ageing Study in India (LASI). Ethical approval for the LASI survey was obtained from the Indian Council of Medical Research (ICMR), and informed consent was secured from all participants by the field survey agencies. This study was additionally approved by the NTU Ethical Review Board (NTU REC-No.: 202407HS010) and adhered to ethical guidelines for the use of secondary data. All analyses were conducted using Python (v3.7). Furthermore, this study followed the guidelines of the Transparent Reporting of a Multivariable Prediction Model for Individual Prognosis or Diagnosis (TRIPOD).

## RESULTS

### Study Population

The final analytical sample included 55, 962 adults aged above 45 years, of whom 41, 025 (73.3%) were aged 45 to 65 years and 14, 937 (26.7%) were 65 years or older. Women comprised 53% of the sample. By caste group, 9, 843 participants (17.6%) were from Scheduled Castes, 9, 439 (16.9%) from Scheduled Tribes, 21, 176 (37.8%) from Other Backward Classes, and 15, 504 (27.7%) from the General category. Overall, 29, 859 individuals (53.4%) reported at least one doctor visit in the past year, 3, 672 (6.5%) had at least one hospitalization, and 4, 422 (7.9%) experienced unmet healthcare needs in the same period.

In examining unmet healthcare needs, notable disparities emerged across caste and monthly per capita expenditure (MPCE) groups. The General caste, comprising 27.7% (15,504) of the sample, accounted for only 21.7% (961) of unmet needs, whereas Scheduled Castes, 17.6% (9,843) of the population, represented 31.2% (1,381) of unmet needs. A similar imbalance appeared in MPCE tiers: the lowest quintile (MPCE 1) made up 20.3% (11,345) of participants but 30.6% (1,352) of unmet needs, while the highest quintile (MPCE 5) accounted for 19.6% (10,969) of participants yet only 13.8% (609) of unmet needs (See Table 1).

**Table 1.**
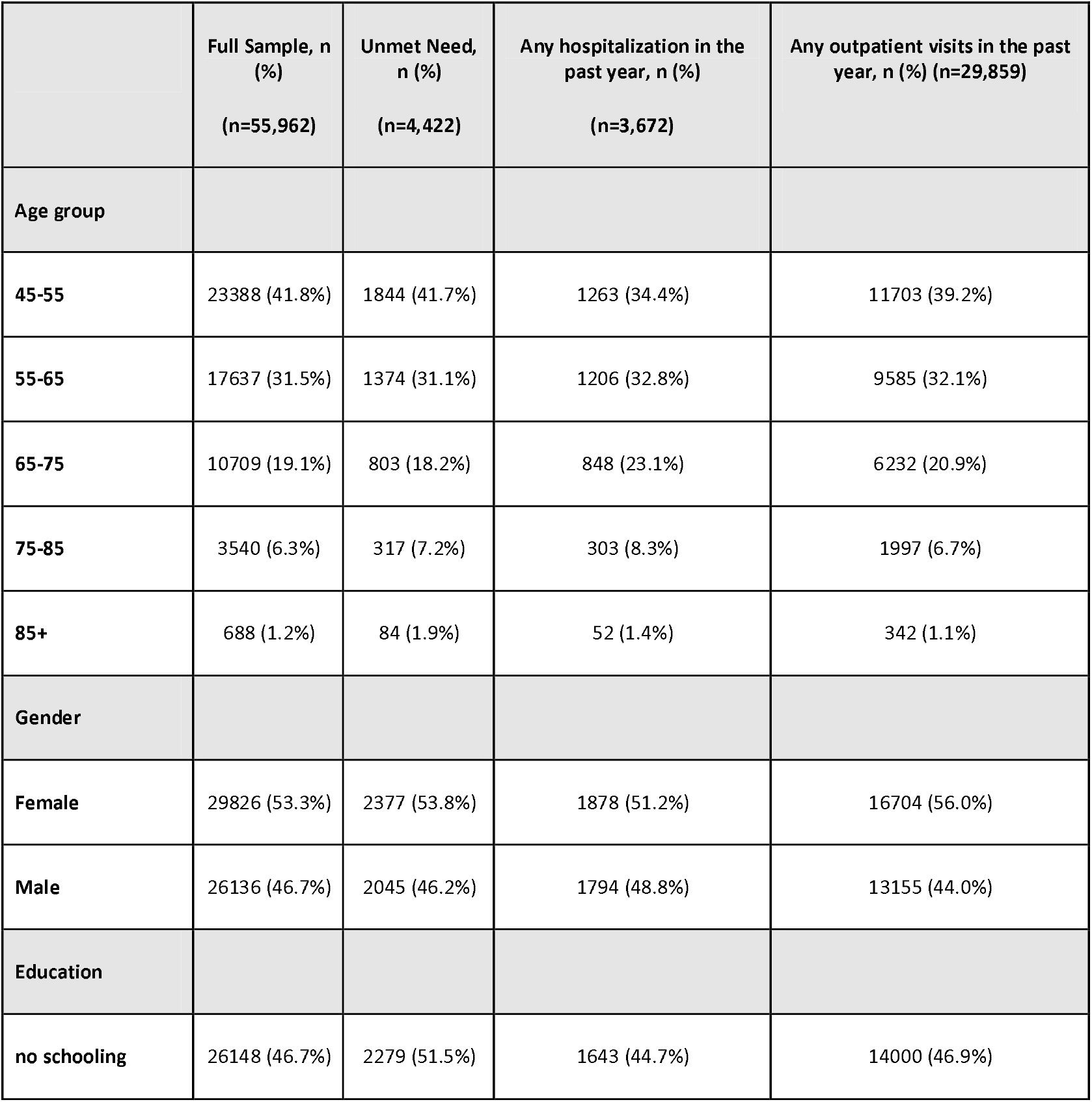
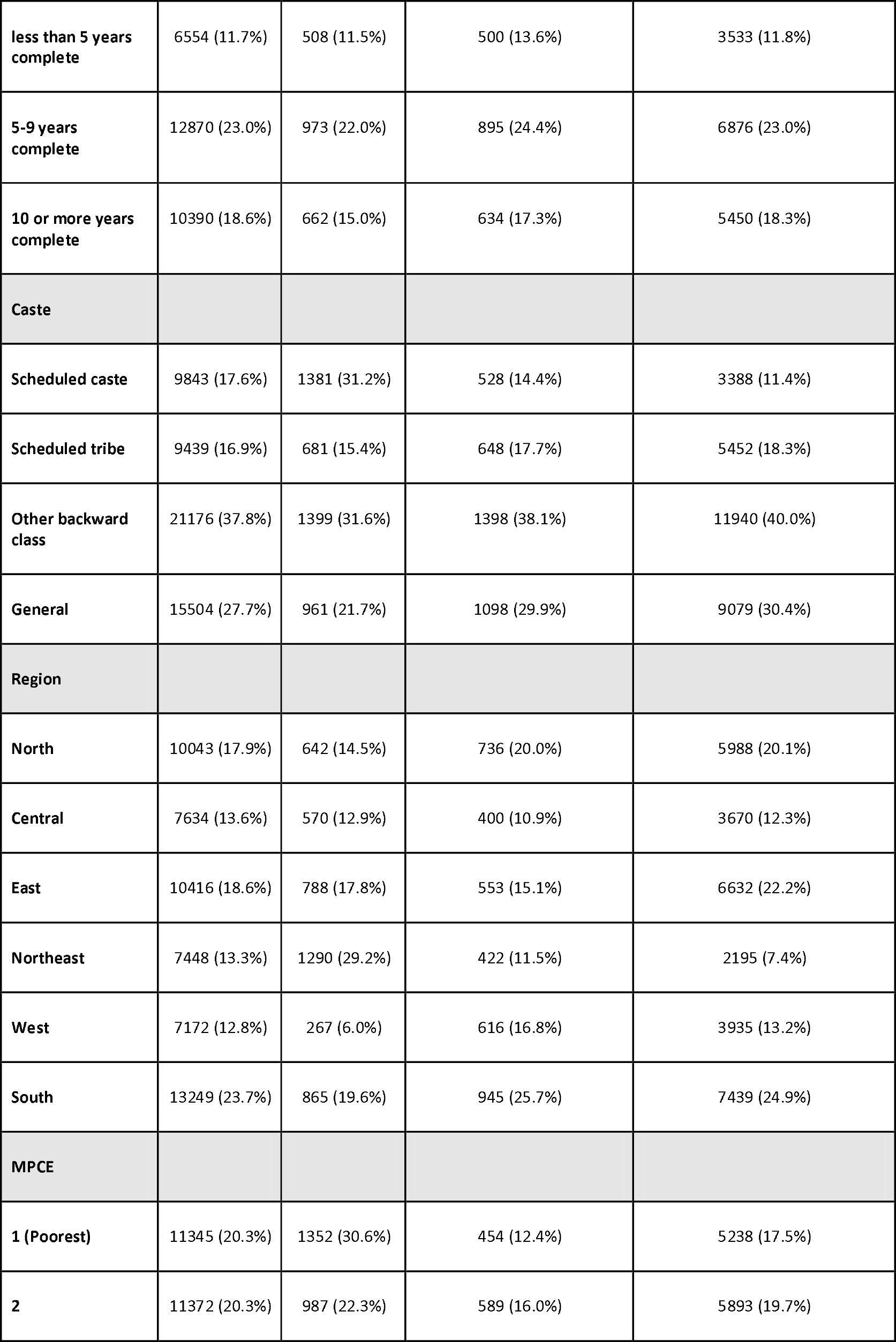
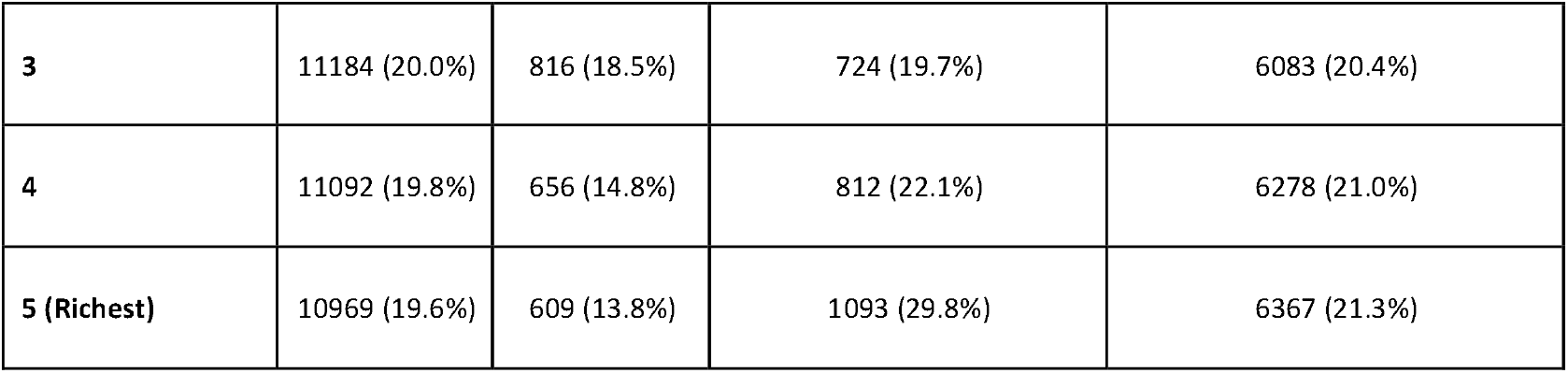
Descriptive summary of outcomes.

### Overall Model Performance

Seven machine learning models—Random Forest, XGBoost, Gradient Boosting, LightGBM, Deep Neural Networks (DNN), Fully Convolutional Networks (FCN), and Logistic Regression—were trained on three key study outcomes: outpatient care utilization, hospitalization, and unmet healthcare needs. Figure 1 (left column) summarizes the area under the receiver operating characteristic curve (AUROC) for each model in the test set. Ensemble methods such as LightGBM, Gradient Boosting, and Random Forest generally outperformed Logistic Regression, DNN, and FCN, with AUROCs ranging from approximately 0.62 to 0.78, depending on the outcome variable. For example, LightGBM achieved an AUROC near 0.78 for unmet healthcare needs, while Logistic Regression and FCN were closer to 0.70. Further details on AUROCs and their 95% CIs can be found in Supplementary Table.

**Figure 1.**
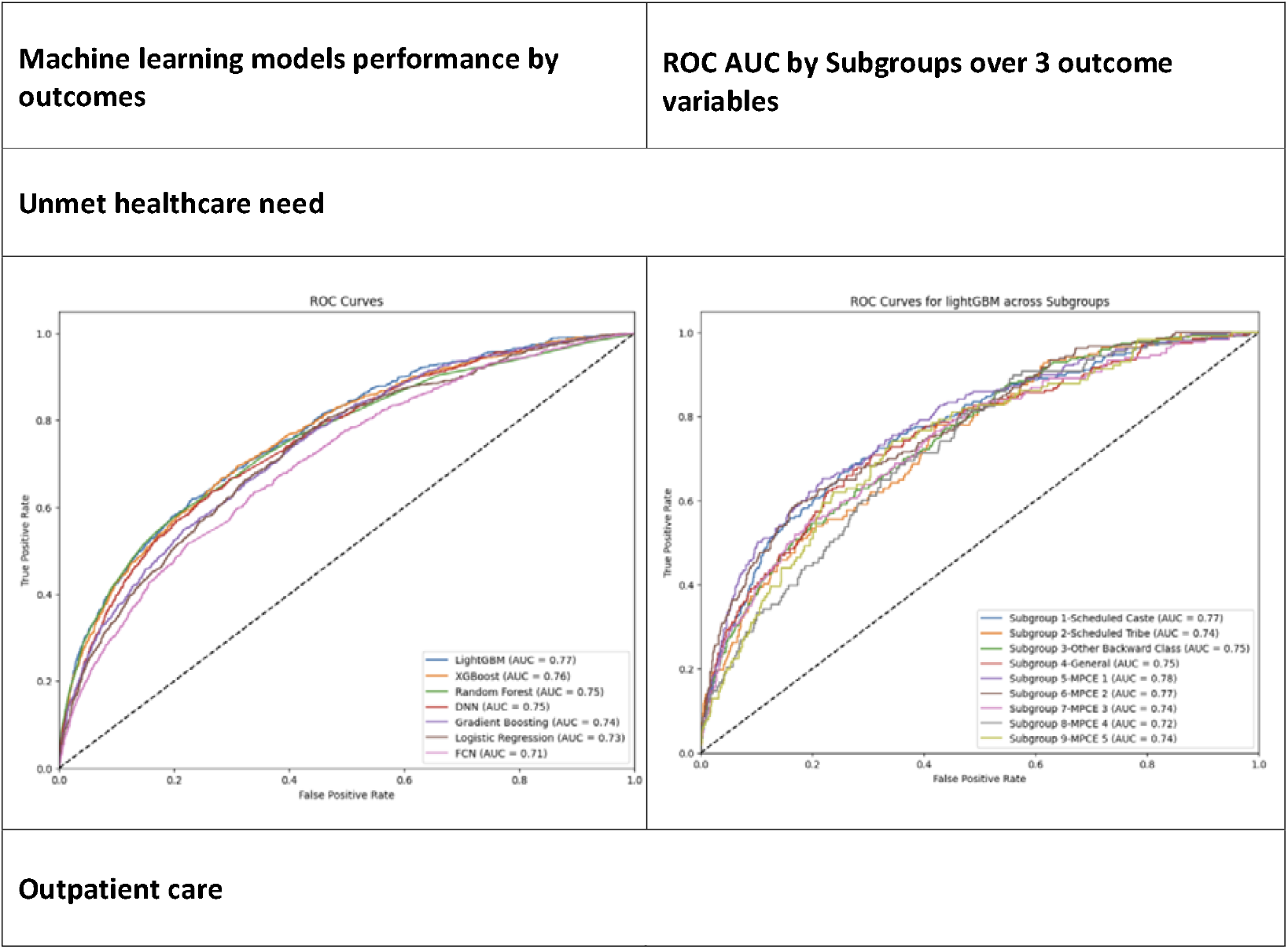
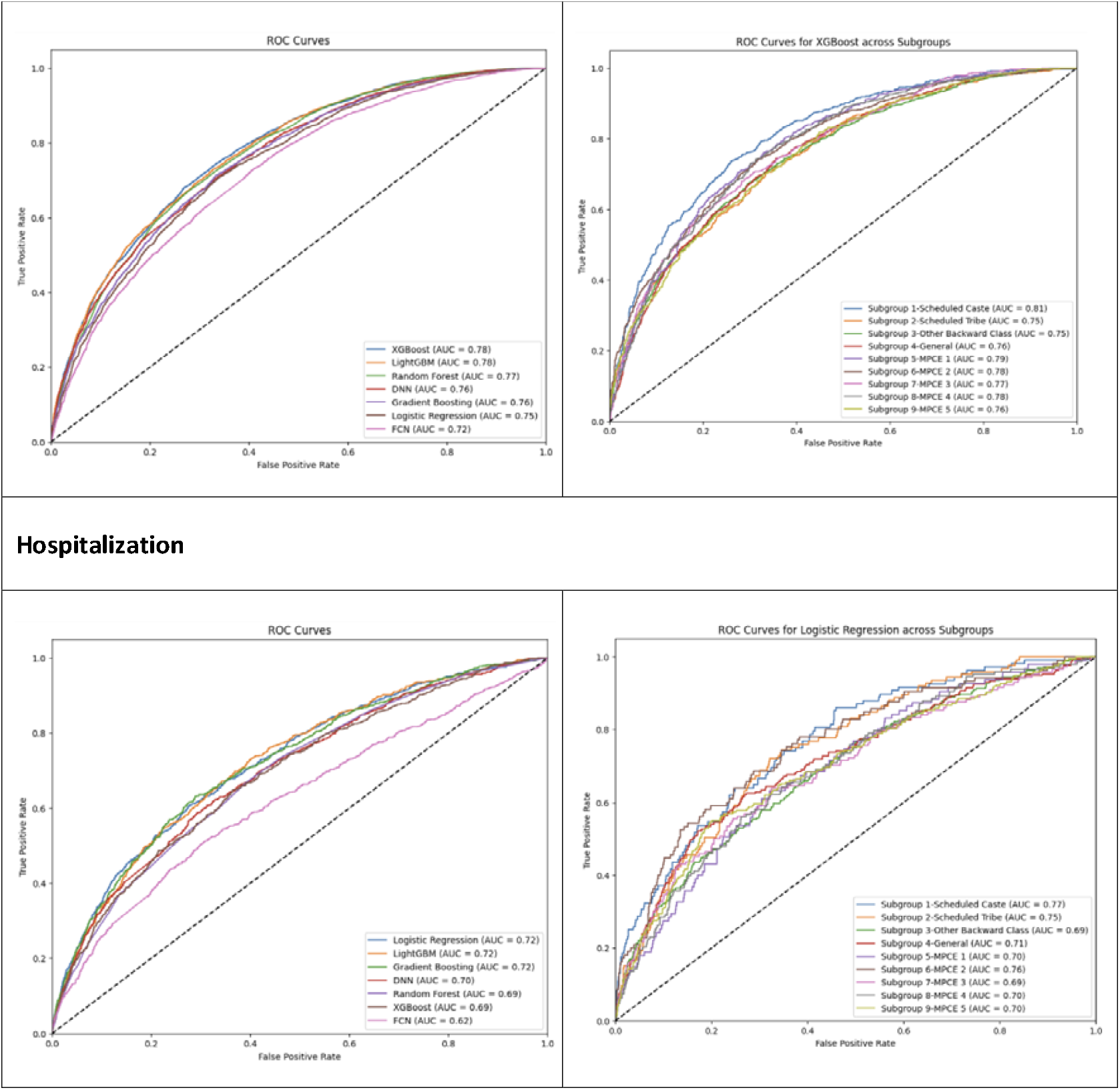
AUROC by Models:

### Evaluating Algorithmic Fairness Across Social Subgroups

We evaluated algorithmic fairness by comparing model performance across caste and socioeconomic subgroups. As shown in Figure 2, predictive accuracy varied substantially across groups. For unmet healthcare needs, the LightGBM model achieved the highest performance in the poorest income quintile (MPCE 1, AUROC = 0.79), compared with AUROC = 0.75 in the wealthiest group (MPCE 5). A similar gradient was observed for outpatient care and hospitalizations, though overall AUROCs were lower.

**Figure 2.**
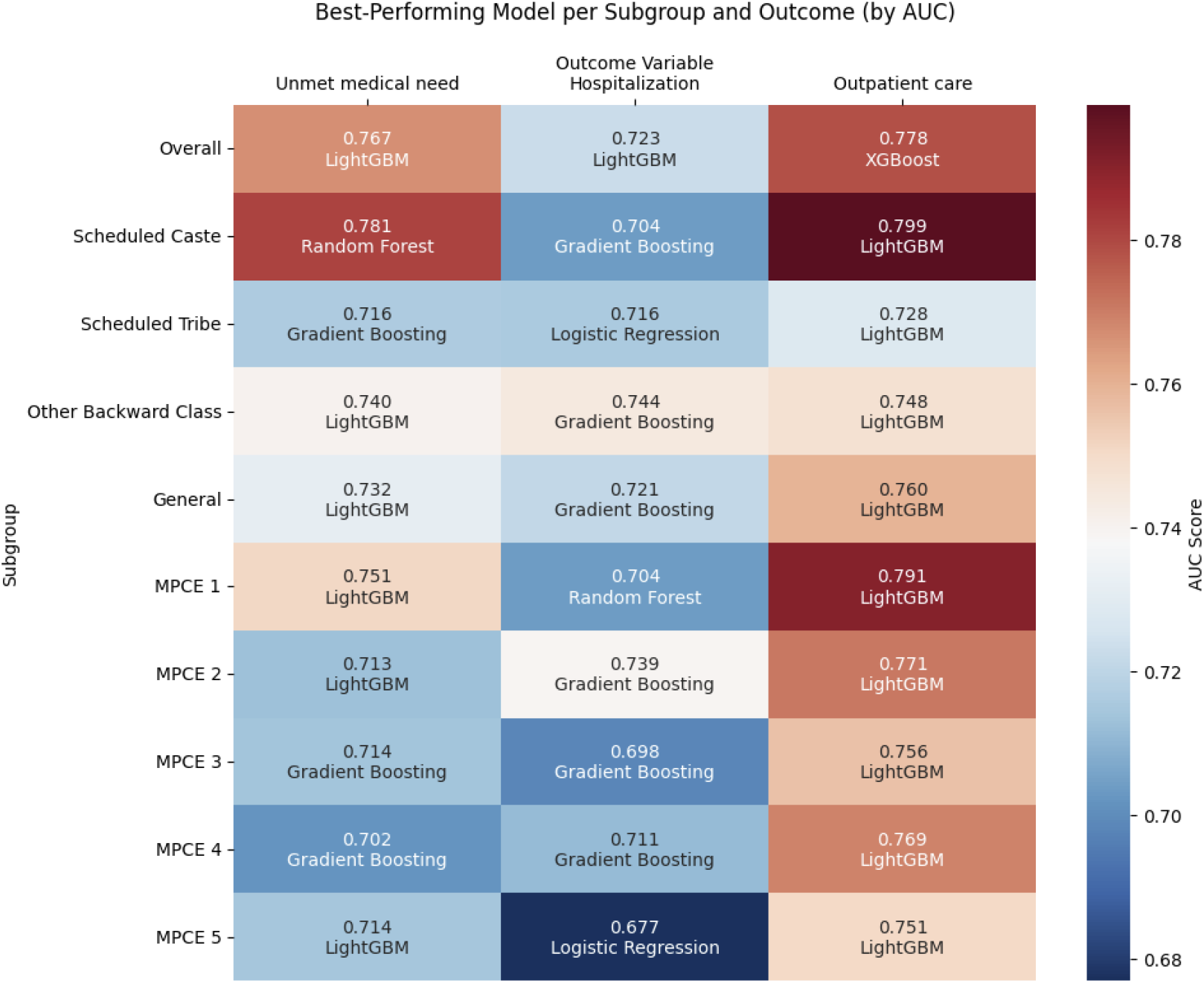
Subgroup best models: Assess model fairness across subgroup.

Across caste groups, the model performed best for Scheduled Castes (AUROC = 0.80) and worst for Scheduled Tribes (AUROC = 0.75) in predicting unmet need. Disparities persisted across all three outcomes, suggesting differential model performance by social position. We also tested two common fairness adjustments. First, models excluding sensitive variables such as caste or income (“neutral models”) showed negligible changes in performance (ΔAUROC < 0.02 across subgroups), indicating that disparities are not resolved by omitting protected features. Second, stratified models trained separately for each subgroup yielded limited improvements: for example, AUROC for unmet need among Scheduled Castes increased marginally from 0.78 to 0.80, while no consistent benefit was observed for other groups.

### Stratified Modeling

Training separate models for each caste and income subgroup yielded limited performance improvements. For most groups, stratified models performed similarly or worse than the pooled model. The most notable gain was for the Scheduled Caste group in predicting unmet need, where AUROC increased from 0.78 (pooled) to 0.80 (stratified). However, stratified models did not improve performance for Scheduled Tribes, Other Backward Classes, or any of the MPCE income quintiles. These findings suggest that group-specific model training does not consistently enhance predictive accuracy, and that fairness challenges may stem from underlying structural disparities rather than algorithmic bias alone (See Figure 2).

### Feature Importance

SHAP analysis identified several key predictors of healthcare use and unmet needs across all three outcomes. Self-rated health consistently ranked as the most influential variable, followed by region, grip strength, and monthly per capita consumption expenditure (MPCE). These factors reflect both individual health status and structural determinants of access. Caste group, living arrangement, and educational attainment also appeared frequently among the top predictors, particularly for unmet need.

For hospitalization, health-related indicators such as self-rated health and physical functioning were dominant. In contrast, predictions of unmet healthcare need were more strongly influenced by socioeconomic and geographic variables, including caste, MPCE, and region—highlighting the role of social disadvantage in limiting access to care. Outpatient care fell in between, shaped by a mix of health and social predictors.

### Incorporating Social and Health Determinants

The inclusion of additional social and health determinants was associated with improved accuracy. As shown in Table 2, a model trained solely on demographic features (D)—such as age, gender, education, and region—often achieved an AUROC around 0.60. Adding social variables (D+S), which include factors like household consumption expenditure (MPCE), working status, and caste, and further incorporating health indicators (D+S+H), such as hypertension, diabetes, and cognitive scores, consistently raised scores to ≥0.70 across multiple outcomes. For example, LightGBM’s hospitalization prediction increased from 0.57 (D only) to 0.70 (D+S+H). Similarly, creating “neutral models” by excluding subgroup-sensitive factors (e.g., MPCE, caste) did not substantially alter performance across subgroups, indicating that removing these explicit predictors alone is not sufficient to address potential biases.

**Table 2.**
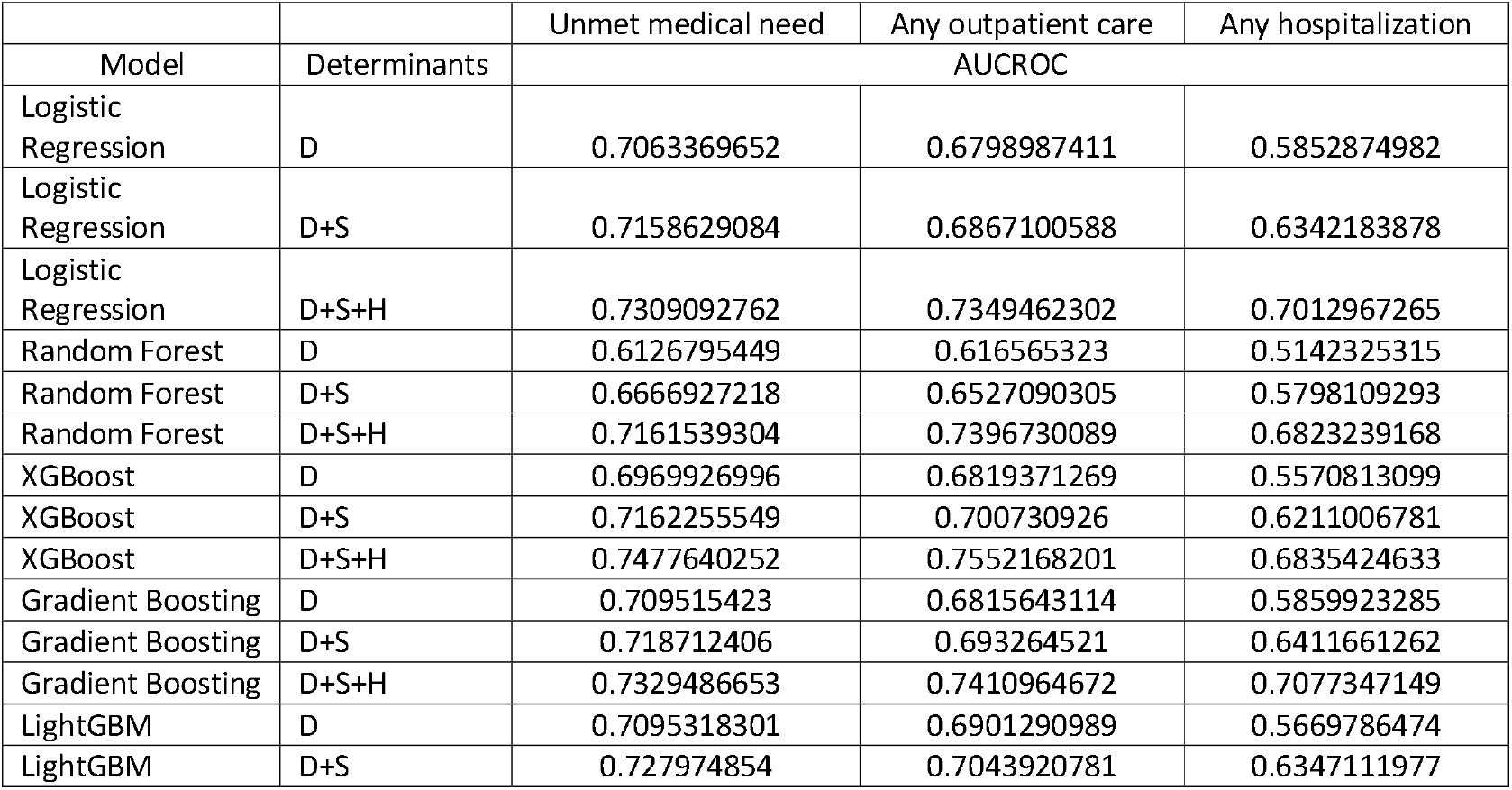
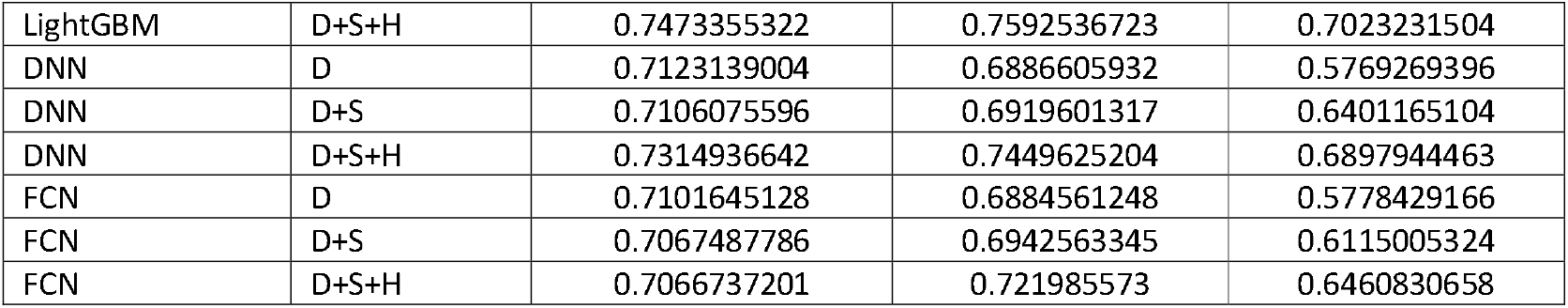
Incorporating socioeconomic and health variables.

|  |  | Unmet medical need | Any outpatient care | Any hospitalization |
| --- | --- | --- | --- | --- |
| Model | Determinants | AUCROC |  |  |
| Logistic Regression | D | 0.7063369652 | 0.6798987411 | 0.5852874982 |
| Logistic Regression | D+S | 0.7158629084 | 0.6867100588 | 0.6342183878 |
| Logistic Regression | D+S+H | 0.7309092762 | 0.7349462302 | 0.7012967265 |
| Random Forest | D | 0.6126795449 | 0.616565323 | 0.5142325315 |
| Random Forest | D+S | 0.6666927218 | 0.6527090305 | 0.5798109293 |
| Random Forest | D+S+H | 0.7161539304 | 0.7396730089 | 0.6823239168 |
| XGBoost | D | 0.6969926996 | 0.6819371269 | 0.5570813099 |
| XGBoost | D+S | 0.7162255549 | 0.700730926 | 0.6211006781 |
| XGBoost | D+S+H | 0.7477640252 | 0.7552168201 | 0.6835424633 |
| Gradient Boosting | D | 0.709515423 | 0.6815643114 | 0.5859923285 |
| Gradient Boosting | D+S | 0.718712406 | 0.693264521 | 0.6411661262 |
| Gradient Boosting | D+S+H | 0.7329486653 | 0.7410964672 | 0.7077347149 |
| LightGBM | D | 0.7095318301 | 0.6901290989 | 0.5669786474 |
| LightGBM | D+S | 0.727974854 | 0.7043920781 | 0.6347111977 |
| LightGBM | D+S+H | 0.7473355322 | 0.7592536723 | 0.7023231504 |
| DNN | D | 0.7123139004 | 0.6886605932 | 0.5769269396 |
| DNN | D+S | 0.7106075596 | 0.6919601317 | 0.6401165104 |
| DNN | D+S+H | 0.7314936642 | 0.7449625204 | 0.6897944463 |
| FCN | D | 0.7101645128 | 0.6884561248 | 0.5778429166 |
| FCN | D+S | 0.7067487786 | 0.6942563345 | 0.6115005324 |
| FCN | D+S+H | 0.7066737201 | 0.721985573 | 0.6460830658 |

**Table 3.**
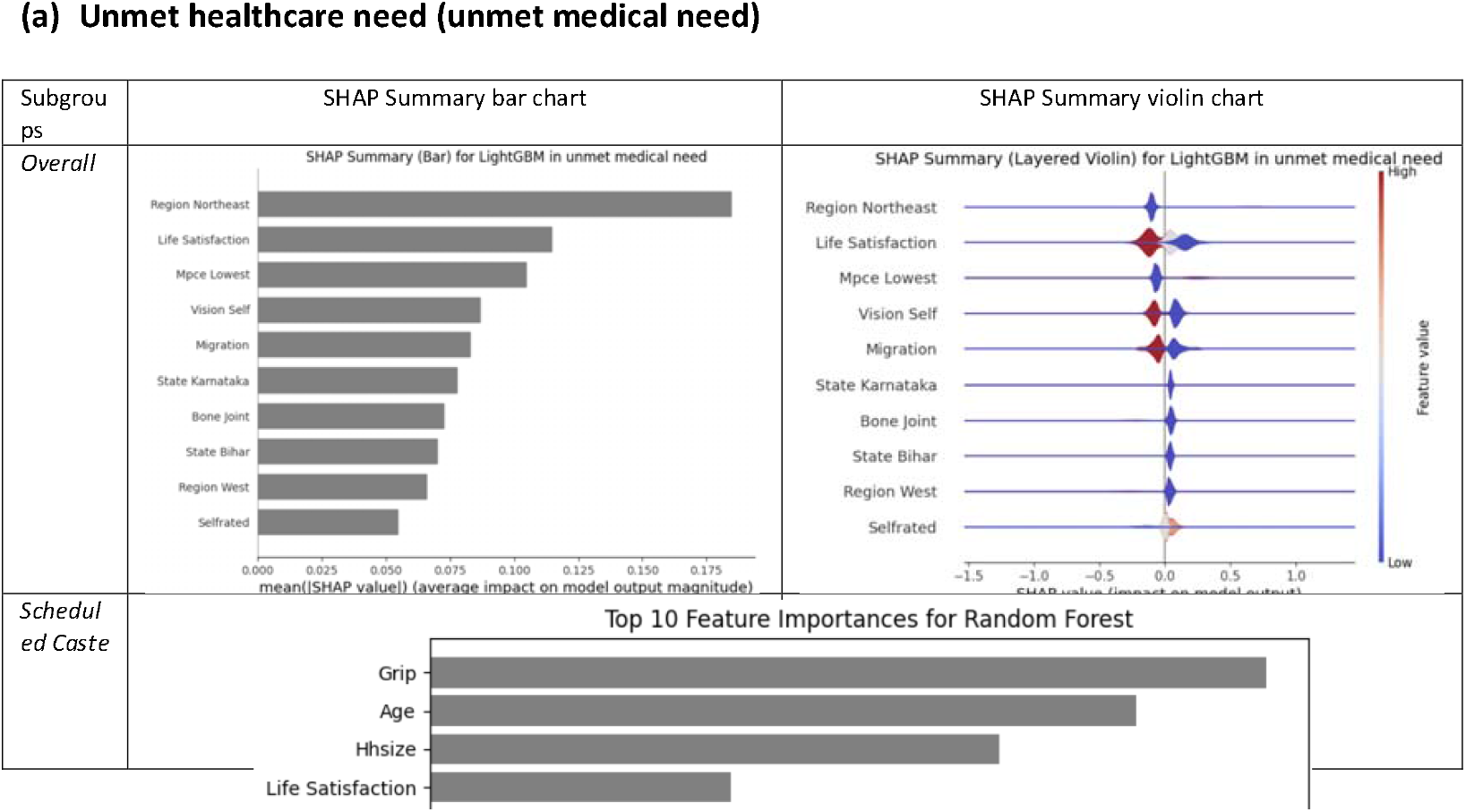
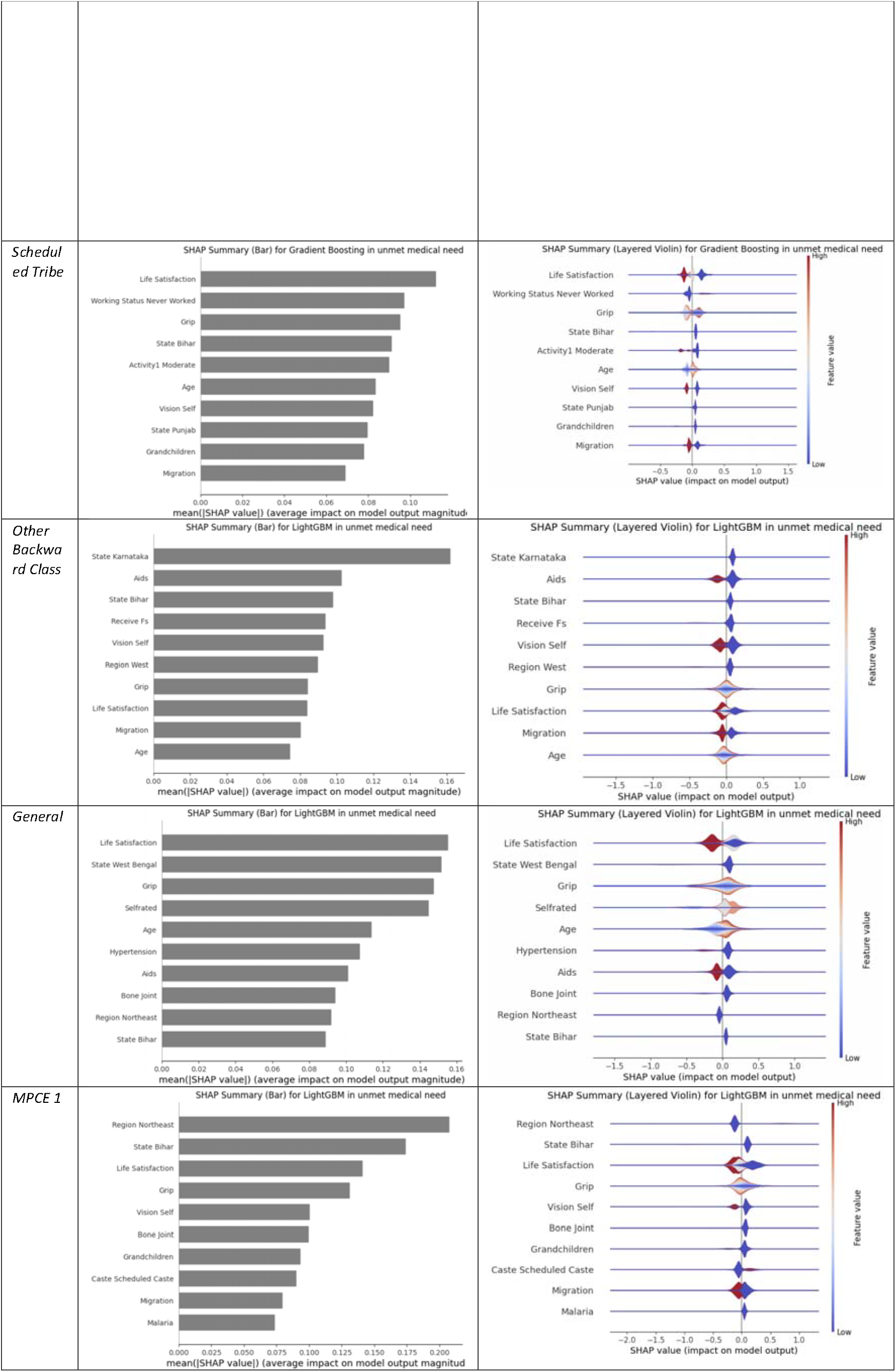
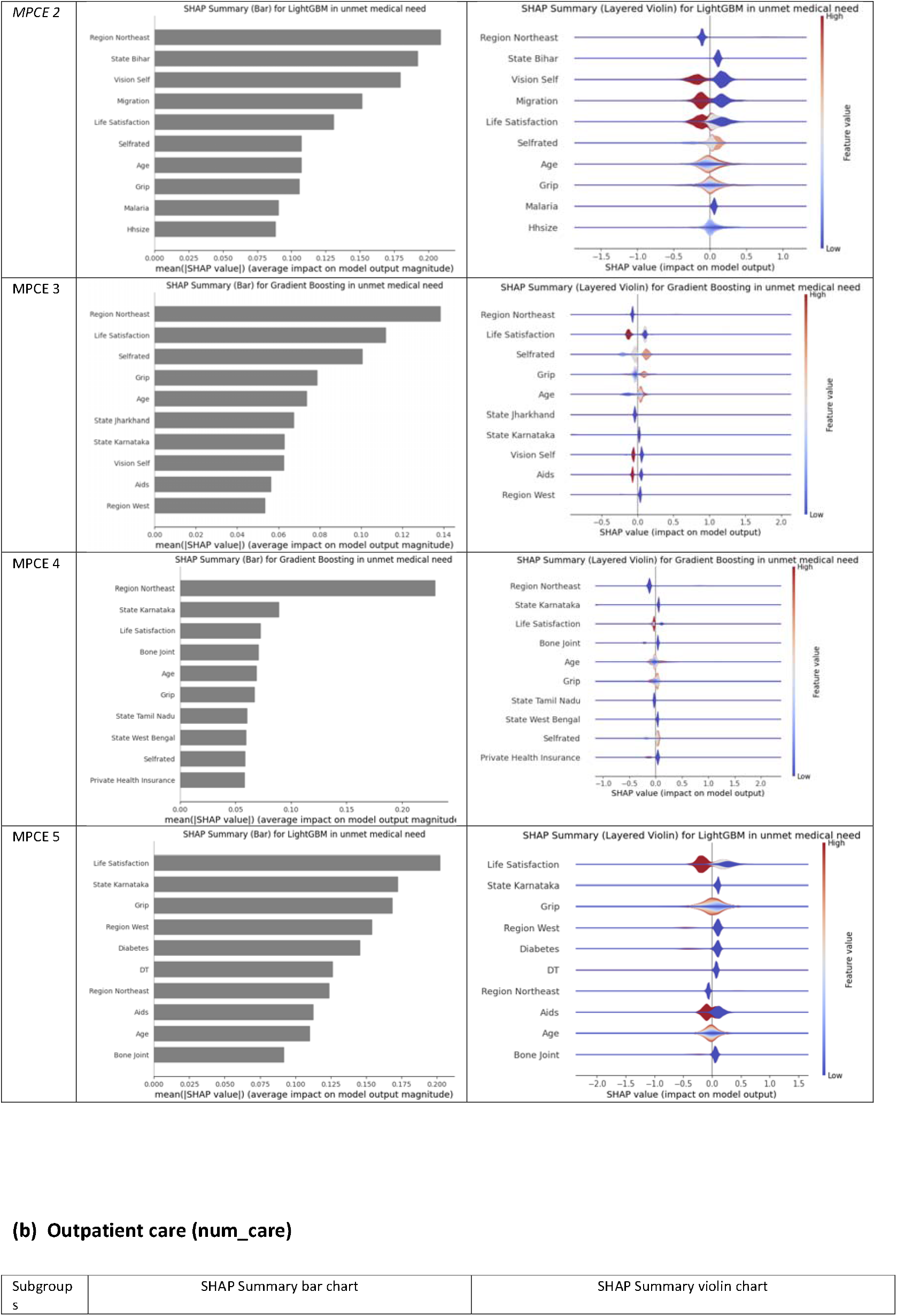
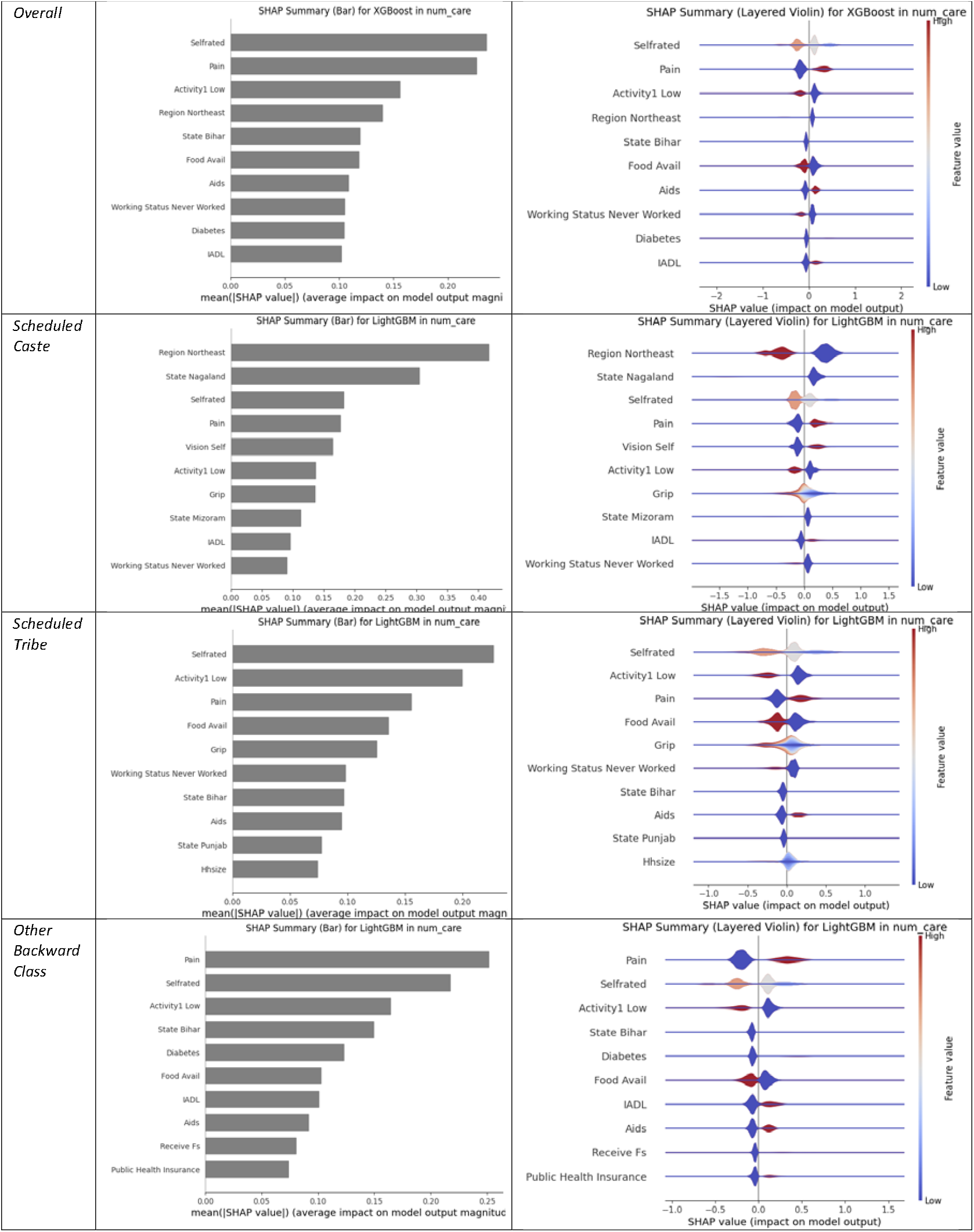
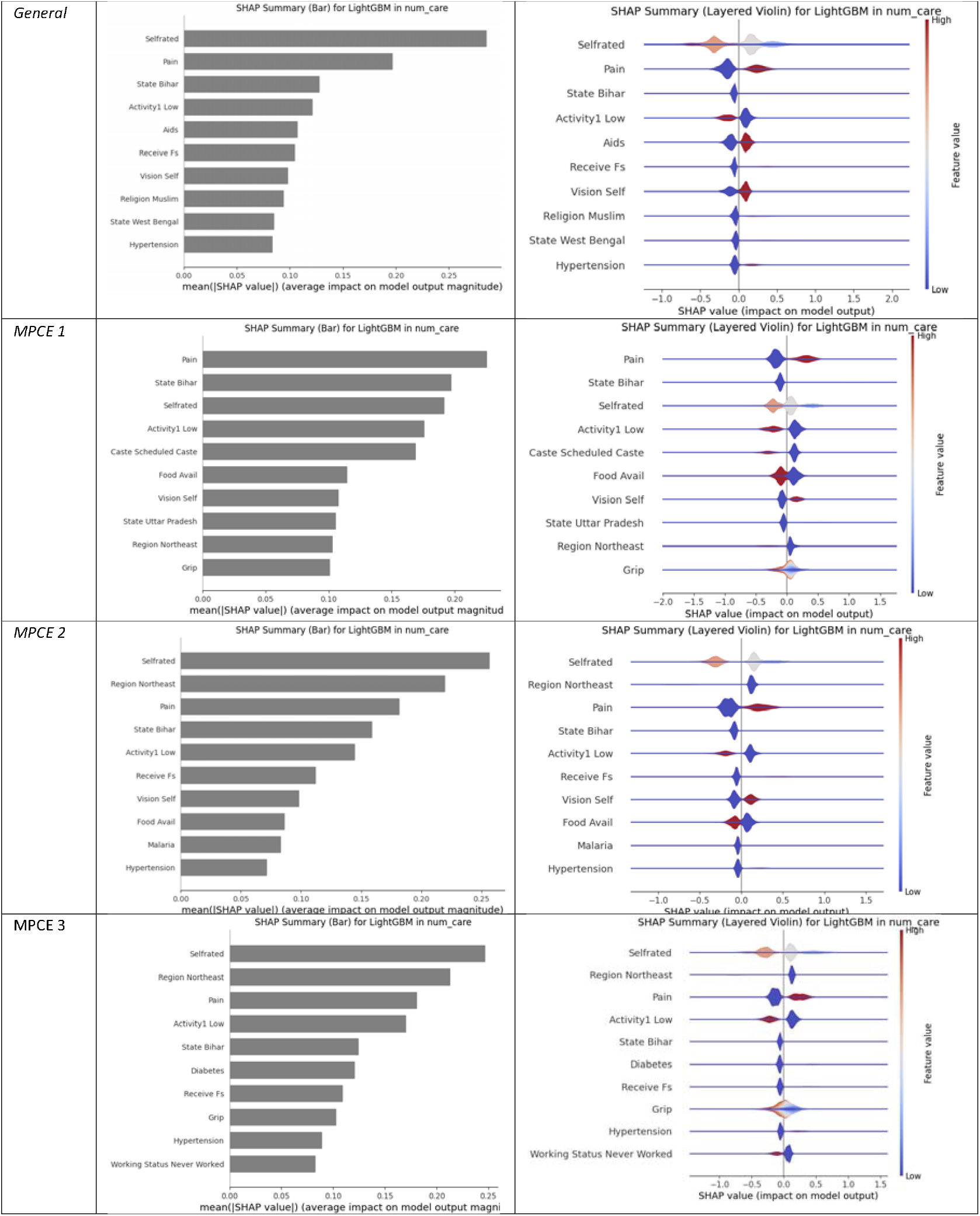
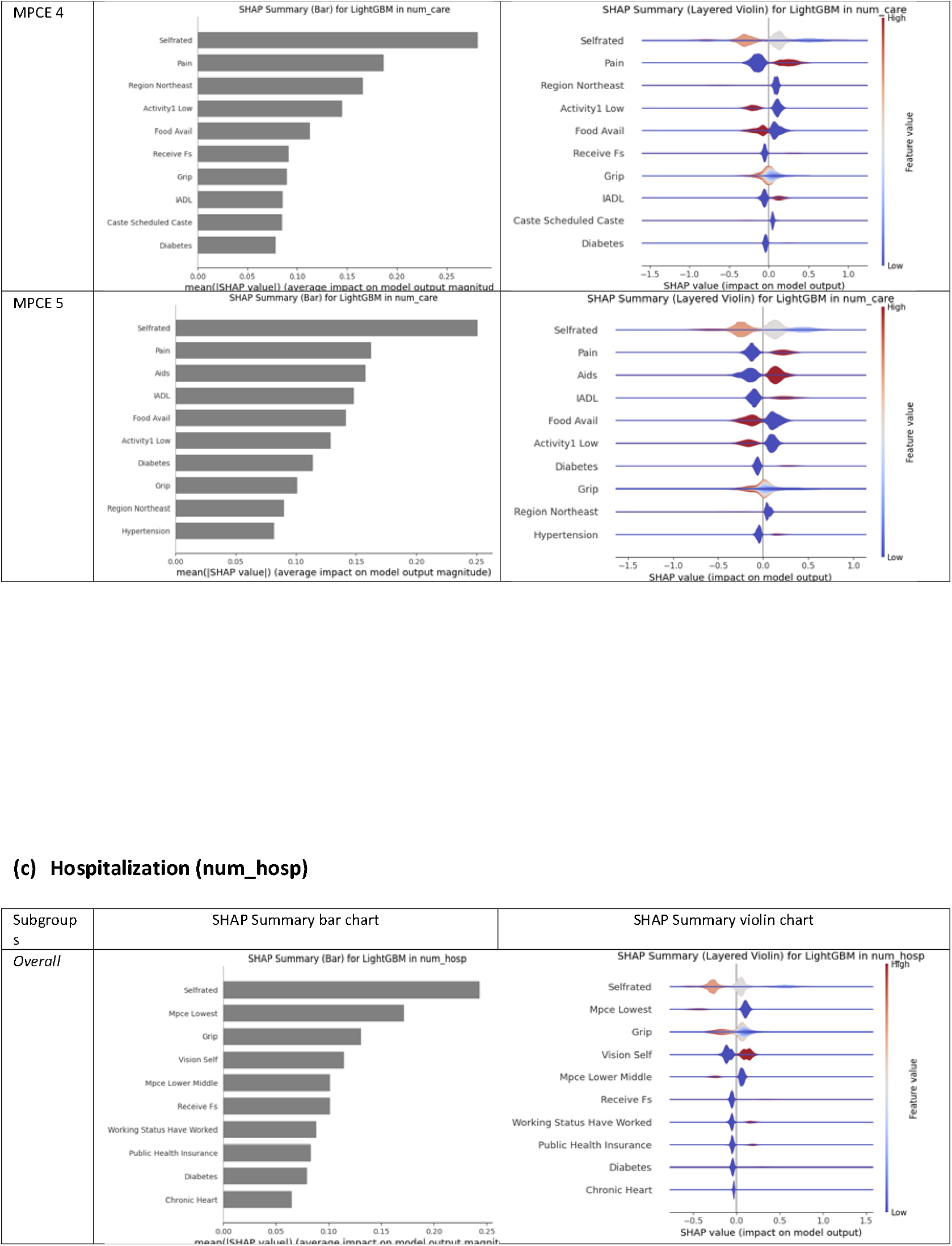
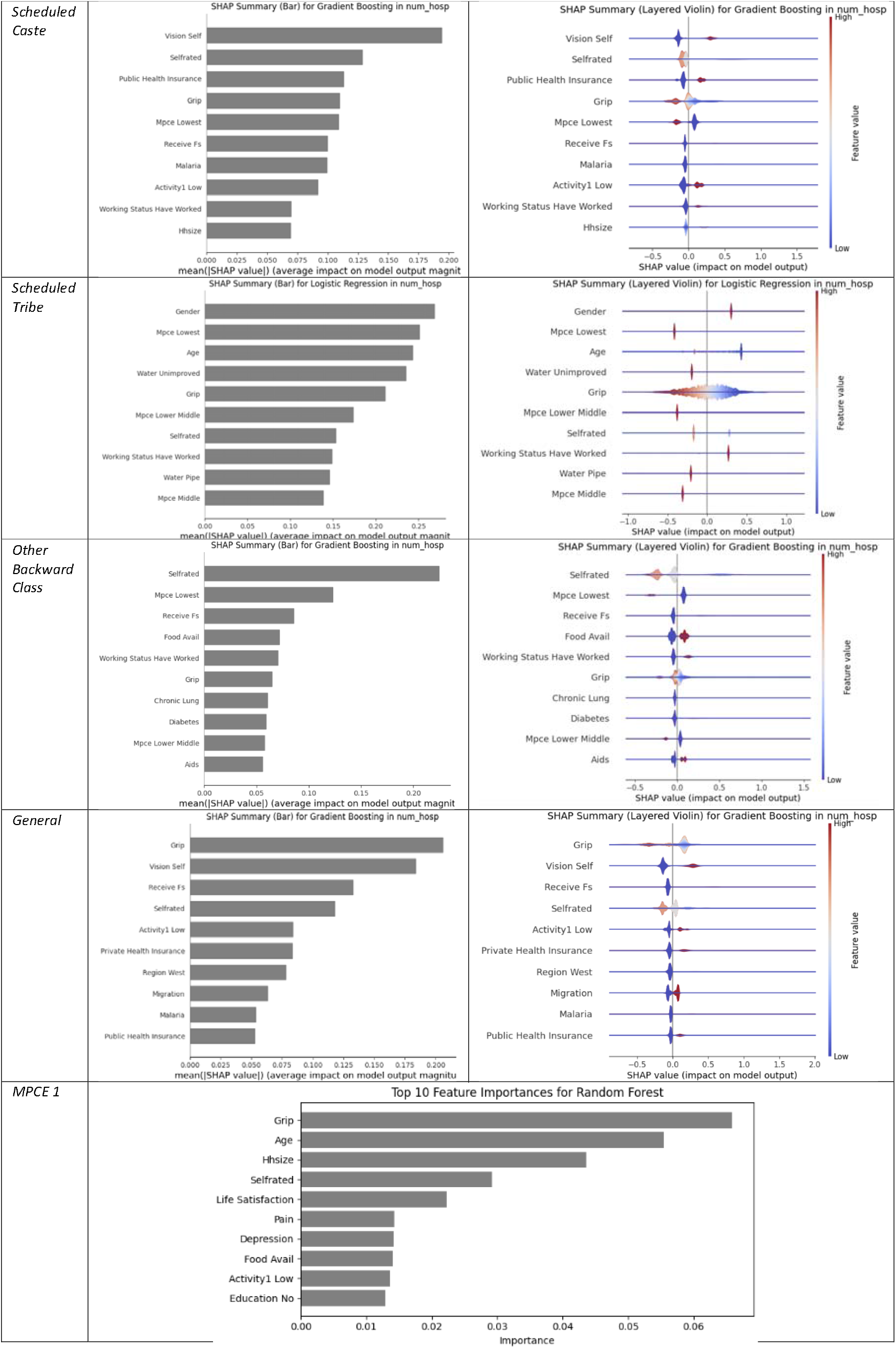
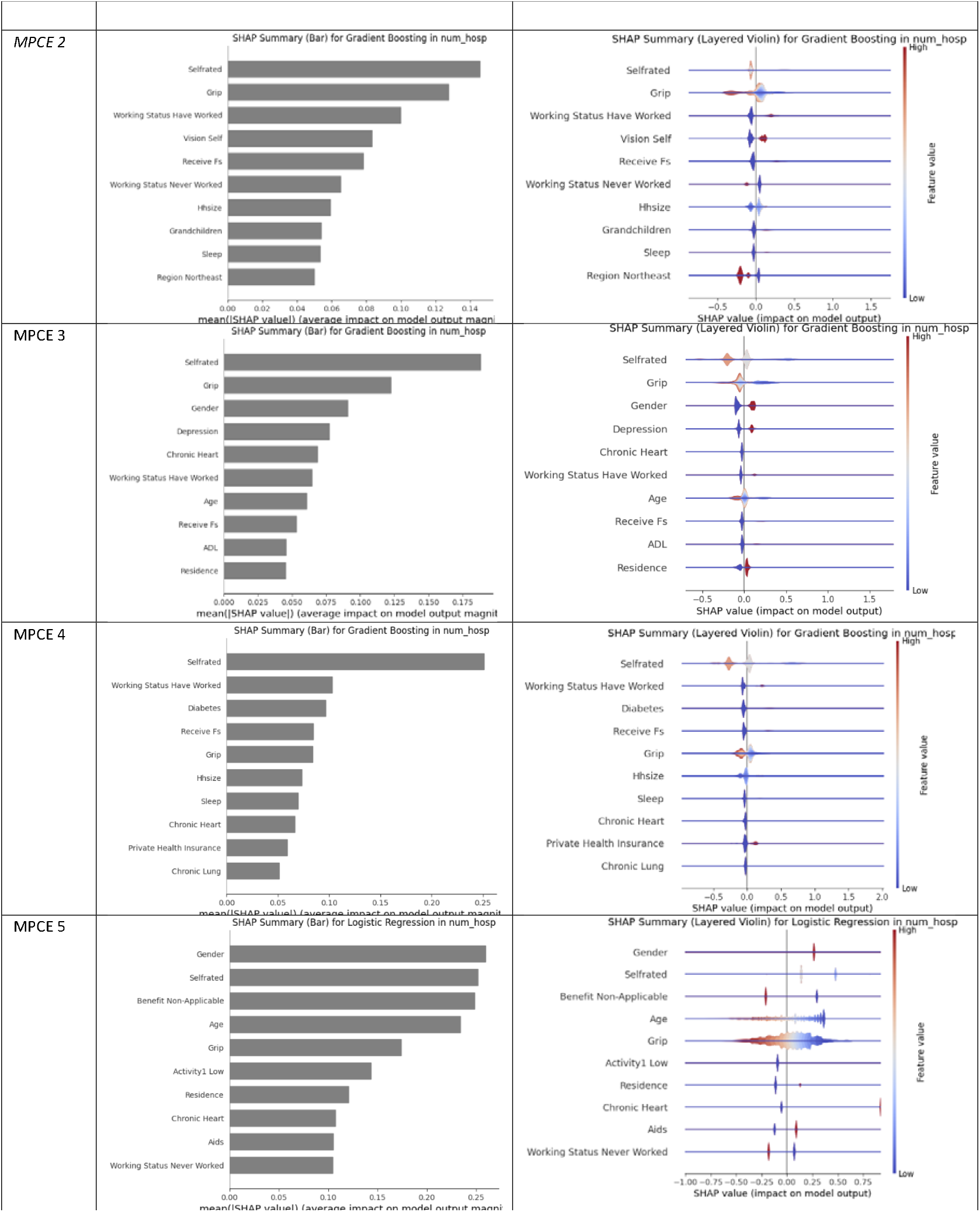
Most Important features by subgroup with best performing models.

### Balancing Techniques

Finally, the synthetic minority over-sampling technique (SMOTE) did not improve performance in most instances and sometimes led to reduced accuracy, suggesting that simple oversampling may not resolve underlying disparities in the data. This outcome indicates the need for additional or more nuanced methods to handle class imbalance and potential subgroup biases.

## DISCUSSION

The study findings highlight that machine learning models, particularly ensemble methods like LightGBM and Gradient Boosting, can outperform traditional approaches in predicting healthcare utilization and unmet needs among older adults in India. However, model performance differs considerably across caste and socioeconomic subgroups, suggesting potential algorithmic bias and underscoring the importance of comprehensive fairness assessments. While removing sensitive attributes (e.g., caste, MPCE) or training subgroup-specific models offered only mixed improvements, incorporating a broader range of social and health determinants substantially increased the models’ predictive power from approximately 0.60 to around 0.70 or higher in AUROC.

SHAP analysis revealed which variables the model relied on most when making predictions. Features such as self-rated health, region, and grip strength strongly influenced model output across outcomes. Importantly, social variables like caste and income also ranked highly—particularly for unmet need—indicating that the model captured patterns of disadvantage embedded in the data. While SHAP values are not causal explanations, they highlight which factors shaped the model’s predictions and point to the importance of including structural determinants in fairness-aware predictive modeling.

Several strategies are proposed in the literature to mitigate algorithmic bias at different stages of the ML pipeline, including data augmentation, adversarial learning, and group-specific threshold adjustments^16, 24^. Our efforts to remove caste and MPCE or to apply SMOTE revealed that no single technique universally corrects subgroup performance gaps, aligning with ongoing debates regarding the lack of consensus on standard bias-mitigation methods.

While strategies such as data augmentation, adversarial learning, and group-specific threshold adjustments have been proposed to mitigate bias, our attempts indicate that no single technique universally eliminates subgroup performance gaps, which is consistent with other studies. Davoudi et al. found significant differences in fairness metrics among heart failure patients when predicting hospitalization in home healthcare, with certain subpopulations consistently outperforming others^25^. Meng et al. reported that deep learning approaches can amplify demographic biases, resulting in disparate treatment across ethnicity, age, and gender^26^. Integrating social determinants of health improved both accuracy and fairness in heart failure diagnosis, as shown by Li et al.^27^. Our study likewise observed that adding social and health determinant variables enhanced AUROC values while potentially helping identify underrepresented groups. Furthermore, Schuch et al. documented bias in machine learning models for preventive dental care ^11^, reinforcing the notion that model performance can deviate for minority or disadvantaged subgroups unless specifically accounted for.

### Strength and Limitation

This study is the first to assess algorithmic fairness in predicting health outcomes within India’s diverse population, addressing caste-based disparities—a critical yet underexplored area in global ML fairness research. Using a comprehensive dataset with 130 variables and integrating social determinants of health (SDOH), the study improved predictive performance (AUROC from 0.6 to 0.7); however, fairness across subgroups did not improve significantly using Neutral model and Stratified model. It highlights that simply removing variables like “caste” or “MPCE” is insufficient to address biases, as deeper structural inequities persist. By tailoring models through data preprocessing, subgroup analyses, and multidimensional feature integration, the study demonstrates how SDOH can potentially improve model performances.

This study did not incorporate survey weights into the machine learning models due to the lack of consensus on how to apply weighting methodologies in predictive analytics. As a result, the representativeness of our findings may be affected if certain subgroups were under- or over-sampled. Furthermore, our analysis relied on cross-sectional data, limiting inferences about causality or changes in healthcare behaviors over time. Finally, although we included a broad set of demographic, social, and health variables, unmeasured confounders may also contribute to subgroup disparities in model performance.

## POLICY AND RESEARCH IMPLICATION

This study highlights the critical role of fairness-aware machine learning (ML) models in addressing healthcare inequities among diverse socioeconomic and caste subgroups in India. Findings reveal major unmet healthcare needs and uneven service utilization, with subgroup-specific modeling outperforming nationwide approaches by capturing population heterogeneity more effectively. LightGBM was the most effective model but exhibited performance disparities, achieving higher accuracy for Scheduled Castes while underperforming for Scheduled Tribes and affluent populations. Key predictors, including socioeconomic status, self-reported health, region, and grip strength, emphasize the need for incorporating social and health determinants into ML frameworks^13,17^.

Universal health coverage and improving access to healthcare for lower-socioeconomic groups is critical for achieving Sustainable Development Goal 3 (Universal Health Coverage). Progress towards UHC requires appropriate and equitable allocation of resources, which can be enhanced through use of fairness-aware ML algorithms that prioritize transparency, inclusivity, and equity in healthcare decision-making. These models, however, must ensure consistent predictive accuracy across subgroups, addressing systemic disparities while avoiding favoritism.

To support equitable health systems, machine learning models should not only predict risks but inform targeted action. Public health planners could use these models to proactively identify population groups—such as Scheduled Castes or the poorest quintiles—who are at high risk of unmet healthcare needs. This would allow for more precise allocation of community outreach, subsidies, or mobile services. Integrating such equity-focused tools into existing programs like Ayushman Bharat can enhance the efficiency and fairness of health service delivery.

Policymakers should invest in robust data infrastructure and disaggregated data collection that are representative of vulnerable and socially-disadvantaged population subgroups, to support subgroup-specific analyses, reduce biases, and refine resource allocation. Ethical guidelines are essential to govern ML applications, ensuring underserved populations like Scheduled Castes, Scheduled Tribes, and women are prioritized. By embedding fairness-focused ML frameworks into public health policies, India can tackle entrenched health inequities effectively and set a global benchmark for ethical, equitable AI-driven healthcare systems^16,25,26^.

## CONCLUSION

Our study reinforce that predictive accuracy alone is insufficient. To meaningfully reduce disparities, machine learning tools must be integrated into the planning and delivery of services, with explicit safeguards to monitor and address subgroup performance gaps. Ensuring equitable model deployment—supported by disaggregated data and aligned with national coverage goals—can help translate algorithmic insights into fairer health outcomes.

## CONTRIBUTORS

JTL conceived the study. VCSL performed the analyses, wrote the results, and contributed to the discussion section. TKBS wrote the policy implications. VTNL assisted in manuscript editing and contributed to the discussion. SHH was responsible for the methods section and provided assistance in analyses. RA, AP and TPL helped revised and gave suggestions to the manuscript.

## DATA AVAILABILITY

Statistical analysis code in python is available for download at https://github.com/johntayuleeHEPI/LASI-Health-Utilization/tree/main

## ACKNOWLEDGEMENTS

The authors express sincere gratitude to International Institute for Population Sciences, IFLS data. Additionally, we thank our colleagues and advisors for their insightful feedback and support throughout the research process. JTL would like to thank Mount Jade Fellowship from the Ministry of Education in Taiwan.

## Tables and Figures

**Supplementary Table:**
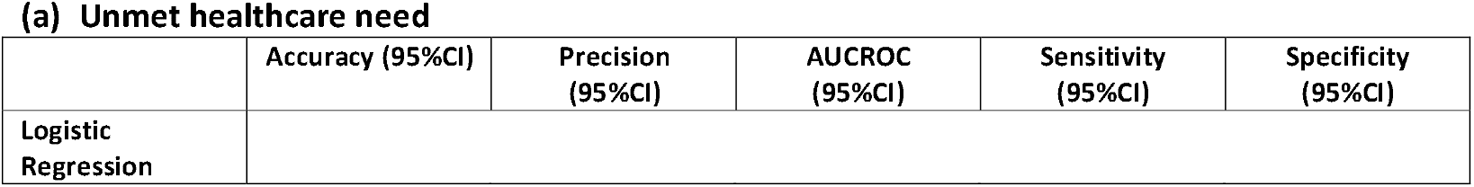
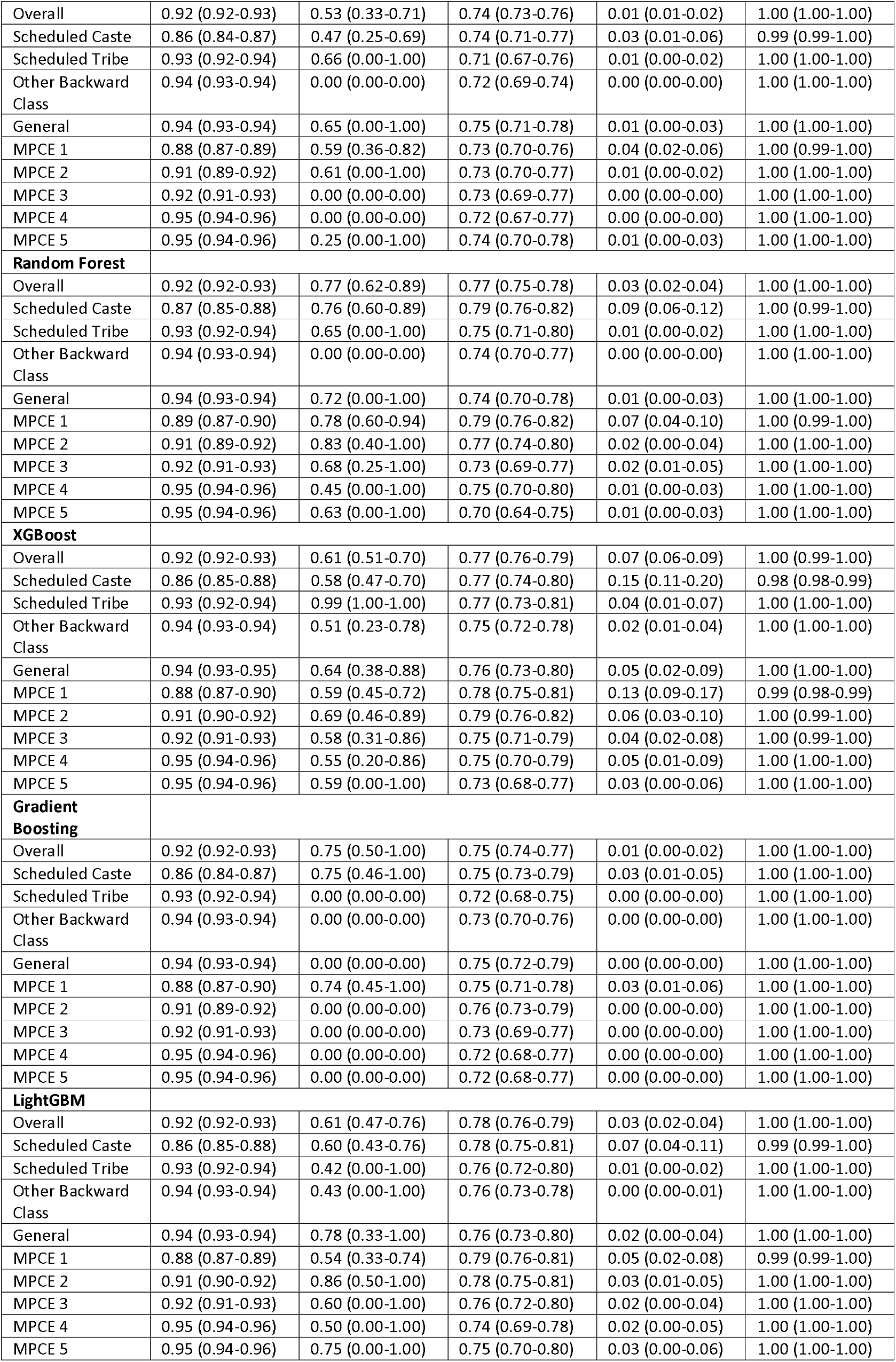
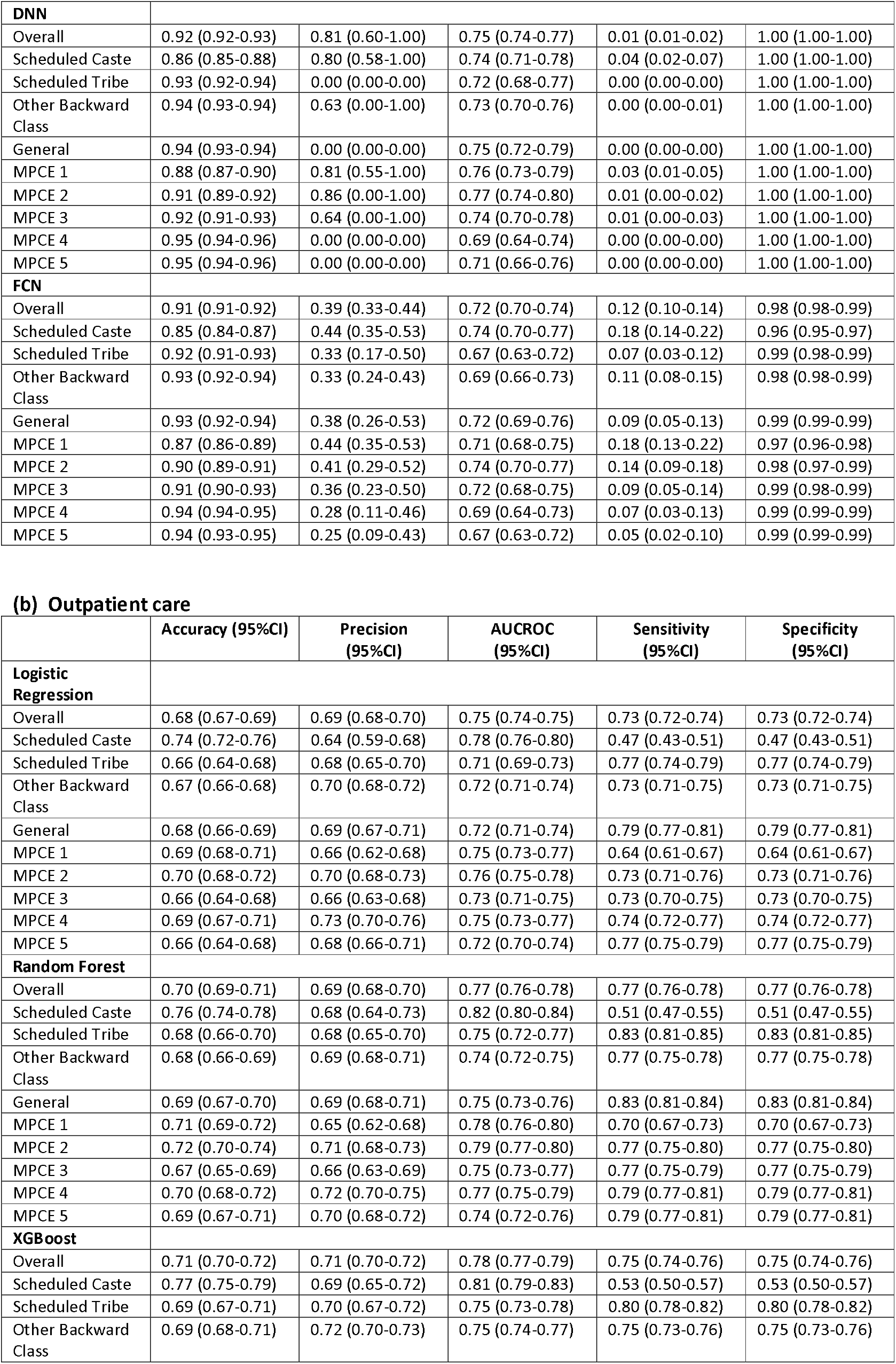
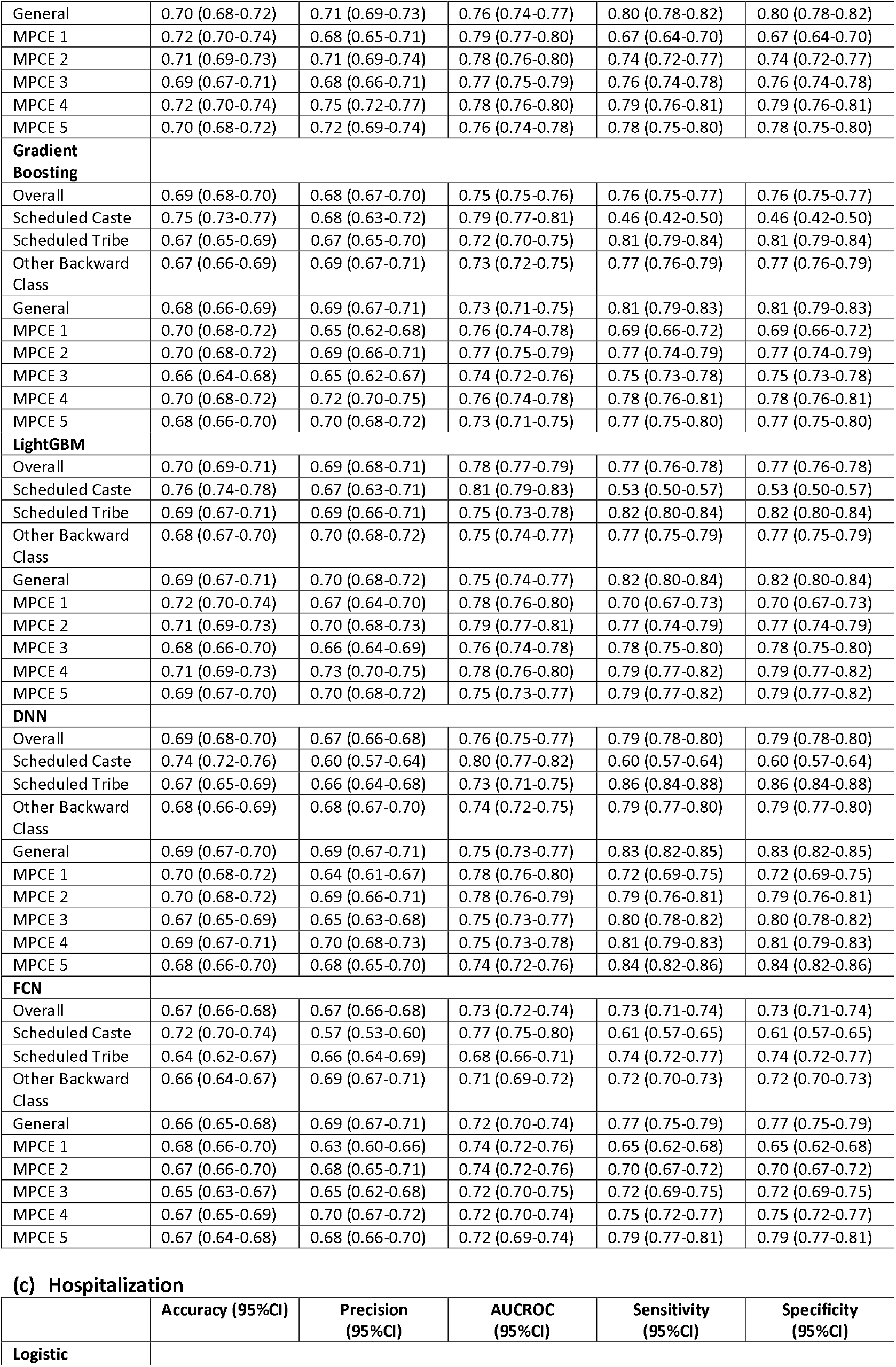
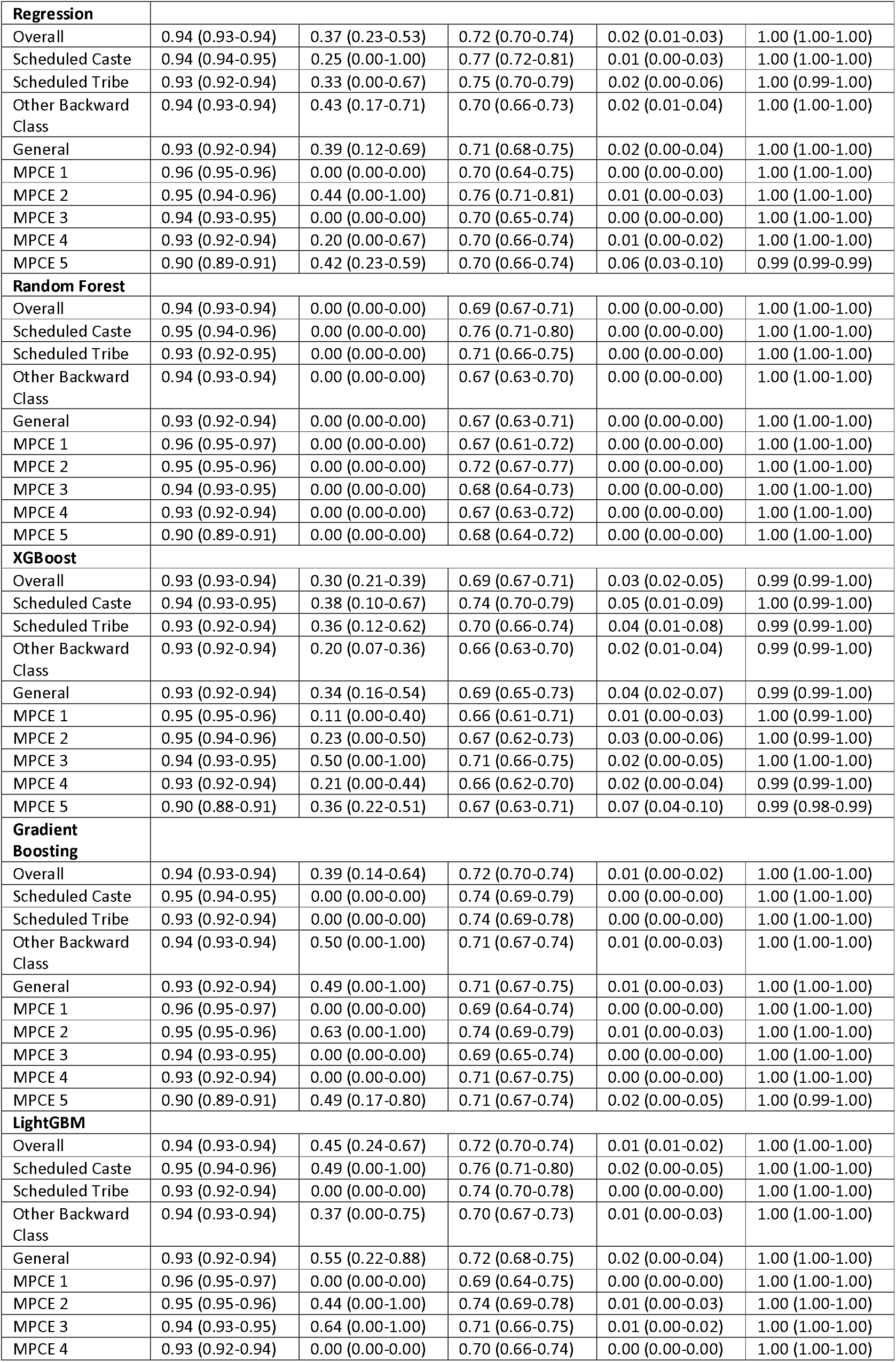
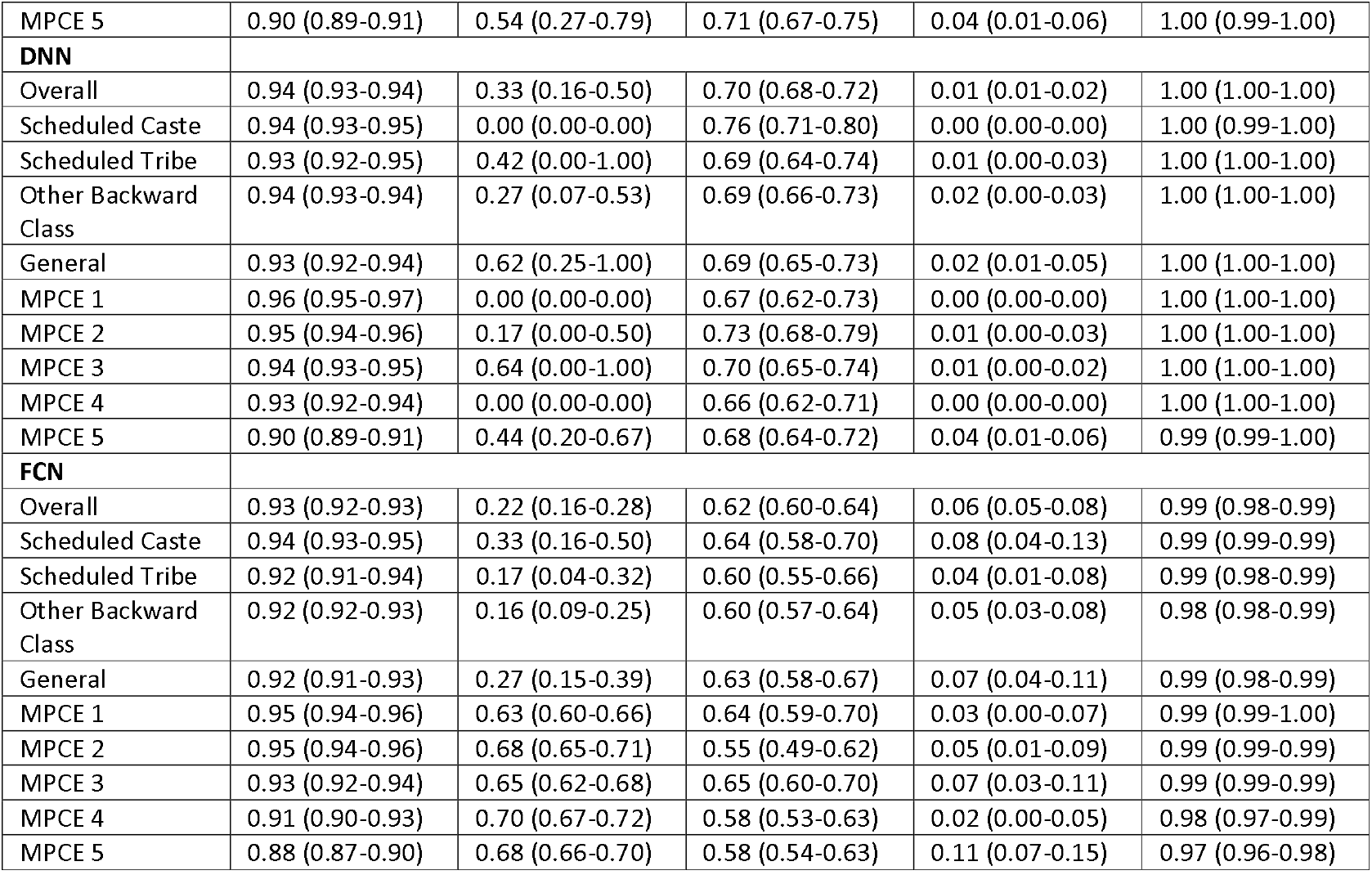
Comparison of Evaluation Metrics Across Different Machine Learning Models.

